# Segmentation of leukoaraiosis on noncontrast head CTs using CT-MRI paired data without human annotation

**DOI:** 10.1101/2024.10.03.24314874

**Authors:** Wi-Sun Ryu, Jae W. Song, Jae-Sung Lim, Ju Hyung Lee, Leonard Sunwoo, Dongmin Kim, Myungjae Lee, Beom Joon Kim

## Abstract

**Background:** Compared to white matter hyperintensity (WMH) on MRI, evaluating leukoaraiosis (LA) on CT can be challenging due to its low contrast-to-noise ratio against white matter and similar attenuation to parenchymal gliosis and edema. We aimed to develop and validate a deep learning algorithm that segments LA using CT-MRI_FLAIR_ paired data from a multicenter, multi-vendor registry from Korea and from a head CT dataset from a US population.

**Methods:** We retrospectively identified CT-MRI_FLAIR_ pairs from a nationwide ischemic stroke registry for training and internal validation (n=482), external testing (n=390), and clinical validation (n=867). Additionally, 100 scans from a US population were collected. WMH on MRI were segmented using previously validated software and the segmentation mask was registered onto the CT scan. Performance was assessed using Dice similarity coefficient (DSC), concordance correlation coefficient (ρ), and Pearson correlation. Predicted LA volumes were analyzed for associations with clinical outcomes.

**Results:** Mean age (SD) for training and external testing datasets were 68.1 (SD 12.7) and 69.2 (SD 13.5) years and 33.2% and 47.9% were female, respectively. External test showed a DSC of 0.527, with ρ values of 0.919 and 0.760 for predicted LA volumes compared to registered LA and WMH volumes on MRI, respectively. In external testing and US datasets, the predicted LA volumes were significantly correlated with Fazekas grade (Pearson correlation coefficient=0.832 and 0.891, respectively). Subgroup analysis demonstrated consistent performance across different CT vendors (ρ ranged from 0.875 to 0.950) and infarct volumes (≤10 mL vs >10 mL; ρ=0.912 and ρ=0.922). In an independent clinical dataset, the predicted LA volumes correlated with age and clinical outcomes after ischemic stroke.

**Conclusion:** Our deep learning algorithm offers a reproducible method for LA segmentation on CT, bridging the gap between CT and MRI assessments in patients with ischemic stroke.

## Introduction

White matter hyperintensities (WMH), also referred to as leukoaraiosis (LA), are the most prevalent brain abnormalities identified on neuroimaging of elderly individuals.^1,2^ The presence and burden of LA are associated with an increased risk of stroke, dementia, depression, and poor outcomes following a stroke.^2–5^ LA is most accurately detected using fluid-attenuated inversion recovery (FLAIR) magnetic resonance (MR) imaging. However, in clinical practice, LA is more frequently identified and progression can be followed by computed tomography (CT) rather than MRI due to the greater availability and accessibility of CT scanners.

Evaluating cerebral LA using CT is more challenging compared to MRI. The hypoattenuation characteristics of LA are less conspicuous against the background of white matter in the presence of gliosis or vasogenic edema on CT scans.^6^ Although several LA scoring systems on head CTs are available,^7^ these systems typically permit only a limited number of ordinal ratings, rely on subjective visual criteria, and have poor associations with quantitative LA volume.^8^ Moreover, the interrater reliability of a visual rating scale using head CTs shows lower agreement (kappa of 0.5∼0.6) compared with brain MRI (kappa around 0.8).^8–10^ Therefore, while visual estimates of LA severity provide valuable prognostic information, they have limited sensitivity as diagnostic tools or markers of disease progression.

Recently, we developed software (JLK-WMH, JLK Inc., Republic of Korea) that automatically segments WMH on FLAIR MRI.^11^ Tested on an external validation dataset comprised of multicenter data (n=6,031), the software showed a high Dice similarity coefficient (DSC) of 0.72.^11^ In the current study, we aimed to develop a deep learning algorithm to automatically segment LA on noncontrast head CT scans. Using a CT-MRI_FLAIR_ paired dataset, we implemented the validated software to segment WMH on FLAIR, then registered the segmentations on the noncontrast head CT, and subsequently trained the algorithm using CT scans without expert annotation. We externally validated this algorithm on an independent testing dataset as well as on a US population dataset. Finally, in a fourth independent clinical dataset, we evaluated the clinical implications of automatically predicted LA volumes in relation to associated risk factors and clinical outcomes after ischemic stroke.

## Methods

### Datasets

This study originated from the Clinical Research Collaboration for Stroke in Korea (CRCS-K), a nationwide web-based registry that records patients with acute ischemic stroke or transient ischemic attack admitted to 20 stroke centers in South Korea.^12–14^ From the imaging substudy, between July 2022 and May 2023, we included 876 patients with available CT-MRI_FLAIR_ paired data for training and internal validation datasets from 4 university hospitals (Figure 1). We then excluded the following patients: duplicated due to recurrent stroke, large (>5mL) infarct core, severe motion artifact on CT or MRI, registration error, incomplete CT slices, contrast enhanced CT, and presence of hemorrhage or brain tumor. Infarct core volumes were measured on diffusion-weighted imaging using verified in-house software.^15,16^ An infarct core volume threshold of >5mL was defined as an exclusion criterion to minimize differences between CT and FLAIR images given the possibility of progression of ischemia and cytotoxic edema with LA on imaging.

**Figure 1.**
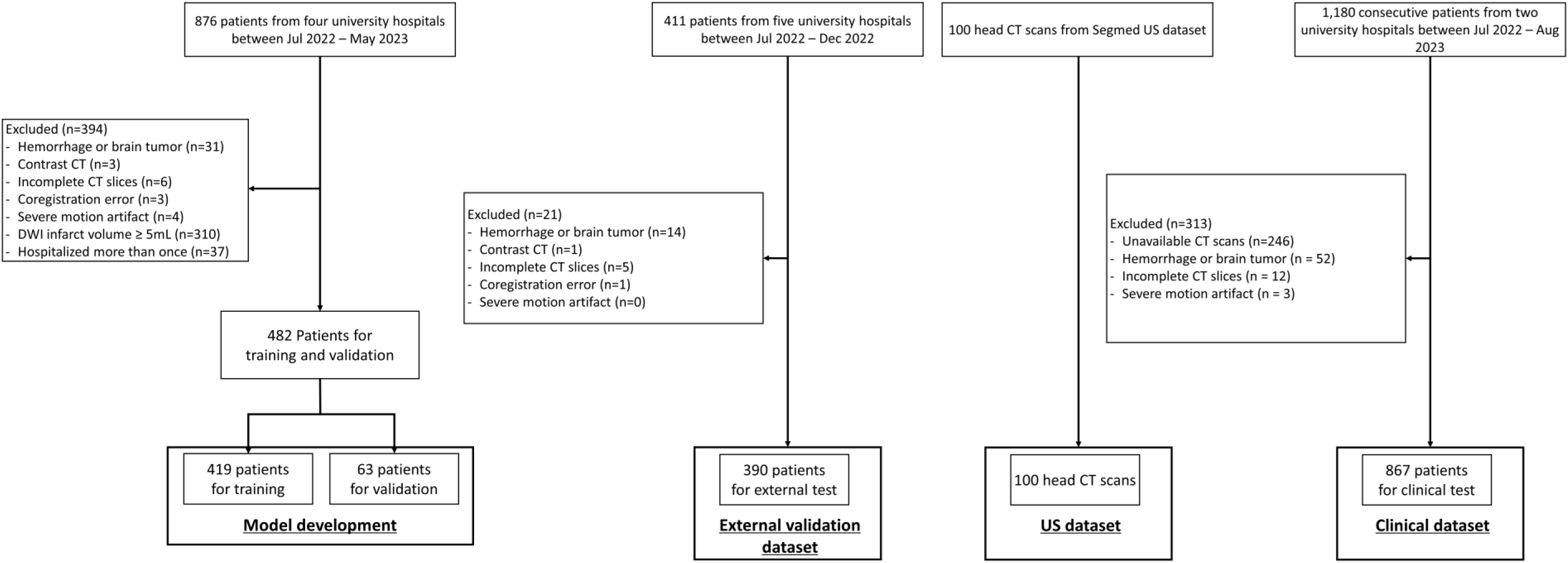
Study flow chart, DWI=diffusion-weighted imaging;

For the external test dataset, 411 patients from five university hospitals were identified between July 2022 and December 2022; these cases did not overlap with those in the training dataset. The exclusion criteria were the same as for the training dataset, except duplicate cases due to recurrent stroke and patients with large infarct cores (>5 mL) were included to evaluate the model’s performance in a real-world ischemic stroke dataset.

For the external US population dataset, we acquired 100 noncontrast head CT along with their radiological impressions from Segmed, Inc. (Stanford, CA). All scans had protected health information, except for age and sex, removed from both the reports and DICOM tags. The cases included were drawn from both outpatient and emergency care settings. We filtered the scans based on the following criteria: (1) > 18 years old; (2) unenhanced; (3) without motion artifacts; (4) slice thickness ≥ 1.5 mm; (5) conducted using a standard convolutional kernel; (6) absence of intracranial hemorrhage or large transcortical infarcts; and (7) axial plane.

A clinical validation dataset was curated to evaluate the clinical relevance of automatically measured LA volumes on CT. From two comprehensive stroke centers in Korea (July 2022-August 2023), 1,180 consecutive patients were identified. These institutions did not overlap with the centers from which the training and validation or external test datasets were collected. We excluded patients if: 1) a CT scan was not available 2) presence of hemorrhagic transformation or brain tumor, 3) incomplete CT slices, and 4) severe motion artifact on CT. Using the initially acquired noncontrast head CT scans, we measured LA volumes using the algorithm. Demographic and clinical data were extracted from the prospective stroke registry. Modified Rankin Scale (mRS) scores at 3 months after stroke and admission National Institute of Health Stroke Scale (NIHSS) scores were collected as previously reported.^4,17–19^ The study protocol was approved by the institutional review board of Seoul National University Bundang Hospital (B-2307-841-303) and all subjects or their legal proxies provided a written informed consent.

### Imaging protocols

For the training and internal validation dataset, the most frequent MRI vendor was Philips (n=273, 56.6%; Table S1), followed by GE (n=127, 26.4%) and Siemens (n=82, 17.0%). The magnetic field strength was 3.0 Tesla (n=415, 86.1%) and 1.5 Tesla (n=67, 13.9%). Most patients had a slice thickness of 5 mm (n=446, 92.5%). For noncontrast head CT scans, the most frequent CT vendor was Siemens (n=262, 54.4%), followed by Philips (n=208, 43.2%), and Canon (n=11, 2.3%). Most patients had a slice thickness of 5 mm (n=432, 89.6%) and underwent CT scans with a kVp of 120 (n=443, 91.9%).

In the external testing dataset, the most frequent MRI vendor was Philips (n=325, 83.3%), followed by Siemens (n=39, 10.0%) and GE (n=24, 6.2%). The magnetic field strength was 3.0 Tesla (n=369, 94.6%) and 1.5 Tesla (n=20, 5.1%). Most patients had a slice thickness of 5 mm (n=287, 73.6%). For noncontrast head CT scans, the most frequent CT vendor was Siemens (n=185, 47.4%), followed by GE (n=91, 23.3%), Philips (n=81, 20.8%), and Canon (n=28, 7.2%). Most patients had a slice thickness of < 5 mm (n=238, 61.0%) and underwent CT scans with a kVp of 120 (n=282, 72.3%).

For the US dataset, the most frequent CT vendor was Siemens (n=54, 54.0%), followed by GE (n=40, 40.0%) and Canon (n=5, 5.0%). Half of the patients had a slice thickness of 5 mm (n=50, 50.0%), followed by 2.5 mm (n=38, 38.0%). Additional details on imaging parameters are provided in Table S1.

### Data preprocessing and preparation

The CT images were first resampled to a target pixel spacing of x=0.41mm, y=0.41mm, and z=5mm to achieve uniform voxel spacing across the dataset. The target spacing was determined by the median voxel size in the given training dataset. Intensity normalization was then applied, which included clipping the Hounsfield Unit (HU) values to a range between −1000 and 1000 HU, covering most anatomical structures relevant for diagnosis, and z-score normalization where the mean and standard deviation of the intensities were used to standardize the images. Cropping and padding were done by automatically detecting the region of interest (ROI) by applying a bounding box around the foreground of the images. If necessary, padding was added to ensure that all images conformed to a minimum size requirement, facilitating batch processing during training.

No human interactions were involved in the process of ground truth generation for CT images. We first generated WMH masks on FLAIR images utilizing validated software,^11^ and applied non-rigid registration using a statistical deformation model^20^ from FLAIR MRI to CT to transform the WMH mask. After registration, the LA mask on CT was thresholded at a probability of 0. Skull-stripping was applied to both FLAIR and CT images beforehand for better alignment of image features.

### Deep learning algorithms: nnUNet framework

We employed the nnUNet framework^21^ and thus used nnUNet architecture in a 2D configuration for deep learning. The 2D nnUNet adopts a similar architecture to the 2D UNet, consisting of an encoder-decoder structure with skip connections.^22^ The encoder path comprises multiple convolutional blocks, each consisting of two 3×3 convolutional layers followed by a rectified linear unit (ReLU) activation and a 2×2 max-pooling layer with a stride of 2. The decoder path mirrors the encoder path in reverse manner. Each decoder block consists of an upsampling layer followed by two 3×3 convolutional layers and a ReLU activation. Between each encoder and decoder block, there is a skip connection with the same resolution in the decoder block. After the decoding step, a 1×1 convolutional layer is applied, followed by SoftMax activation to give the final output. For the learning and optimization step, we used the automated settings of the nnUNet framework, using the SGD optimizer with Nesterov momentum, an initial learning rate of 1e-2, momentum of 0.99, and weight decay of 1e-4 for 1,000 epochs. Dice loss combined with Binary Cross Entropy loss was used for the loss function. Data augmentation was applied during training, including random rotations, scaling, elastic deformations, intensity variations, and flipping.

### Segmentation performance evaluation metrics

The following metrics were used to evaluate the predicted LA segmentation against registered LA mask on CT and the original, automated WMH segmentation on FLAIR:

– Coefficient of determination 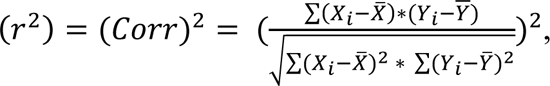
– Concordance Correlation Coefficient 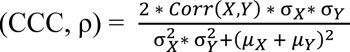

X = predicted LA volume on CT,

Y = registered LA volume on CT or automated WMH volume on FLAIR.

σ = standard deviation, μ = mean

- Dice similarity coefficient 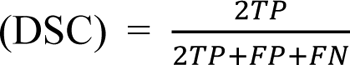, where TP, FP, and FN indicate voxel-level true positive, false positive, and false negative, respectively.

### Experiment and analysis

We implemented the network in Python 3.9.19 using PyTorch 2.3.1. The network (nnUNet) for training was trained on a GeForce RTX A6000 GPU with an 11.8 CUDA version, taking on average 60 seconds per epoch for 14,560/2205 slices (419/63 scans) with the training/validation split. We used a batch size of 12, automatically detected by the nnUNet framework.

After training with the training dataset, the coefficient of determination and concordance correlation coefficient were calculated to evaluate the LA segmentation performance against the external testing dataset. To evaluate the model’s performance against expert manual annotation, an experienced vascular neurologist (W-S. Ryu) with 20 years of experience manually segmented LA on CT scans with referencing FLAIR MRI images, in 40 randomly selected cases from the external testing dataset. In the external test and the US dataset, an expert (W-S. Ryu) visually rated the extent of LA on CT using 4-point Fazekas scale^23^ (none, mild, moderate, and severe) blinded to predicted LA volumes.

### Statistical analysis

Data were presented as the mean±SD or frequency (percentage or interquartile range (IQR)) as appropriate. Baseline characteristics between training and validation datasets versus external test dataset were compared using t-test, rank-sum test, or chi-square test as appropriate. To compare the volumes of predicted LA with registered volumes of LA on CT and WMH volumes on FLAIR images, we utilized the concordance correlation coefficient (CCC: ρ) with 95% confidence intervals (CIs).^24^ To test the relationship between Fazekas grade and predicted LA volumes, we used Pearson correlation coefficient. In the clinical study, associations between demographic and clinical variables and predicted LA volumes were tested using multiple linear regression analyses. The relationship between predicted LA volumes and 3-month mRS score was assessed using multivariable ordinal logistic regression analysis. Because proportional odds assumption was violated, we combined mRS scores 5 and 6 into a single category in the analysis. P < 0.05 was considered statistically significant.

## Results

### Study population

After exclusion, 482 CT-MRI_FLAIR_ paired data from 4 different university hospitals were used for training and internal validation (Figure 1). For the external testing dataset, 390 additional CT-MRI_FLAIR_ paired data from the four stroke centers were included. The mean patient ages for the training/internal validation and external validation datasets were 68.1 (SD 12.7) and 69.2 (SD 13.5) years and 33.2% and 47.4% were female, respectively (Table 1). The median of CT-MRI exam intervals was 3.30 (IQR, 1.04–8.18) and 2.43 (IQR, 1.02–7.04) hours for the internal and external validation datasets, respectively. Median WMH volumes (IQR) on FLAIR were 9.18 mL (4.62–18.5 mL) and 10.23 mL (4.56–22.60 mL), respectively.

**Table 1.** Baseline characteristics of training and validation dataset and external test dataset.

|  | Training and internal validation (n=482) | External test (n=390) | p value |
| --- | --- | --- | --- |
| Age, years (SD) | 68.1 ± 12.7 | 69.4 ± 13.5 | 0.252 |
| Sex, women | 160 (33.2%) | 185 (47.4%) | <0.001 |
| Time interval between CT and FLAIR, hours (IQR) | 3.30 (1.04–8.18) | 2.43 (1.02–7.04) | 0.022 |
| Previous stroke | 88 (21.4 %) | 82 (19.9 %) | 0.565 |
| WMH volume on FLAIR, mL (IQR) | 9.18 (4.62–18.5) | 10.23 (4.56–22.6) | 0.091 |
| Infarct volume on DWI, mL (IQR) | 0.86 (0.16–2.37) | 1.68 (0.39–8.25) | <0.001 |
| Slice thickness of CT |  |  |  |
| < 5mm | 49 (10.1 %) | 238 (61.0 %) |  |
| 5mm | 432 (89.6 %) | 137 (35.1 %) |  |
| > 5mm | 1 (0.21 %) | 14 (3.59 %) |  |
| CT Vendors<sup>a</sup> |  |  |  |
| GE MEDICAL SYSTEMS | 0 (0.00 %) | 91 (23.3 %) |  |
| SIEMENS | 262 (54.4 %) | 185 (47.4 %) |  |
| Philips | 208 (43.2 %) | 81 (20.8 %) |  |
| TOSHIBA (Canon Medical Systems) | 11 (2.3 %) | 28 (7.2 %) |  |
| MRI Vendors<sup>b</sup> |  |  |  |
| GE MEDICAL SYSTEMS | 127 (26.4 %) | 24 (6.2 %) |  |
| SIEMENS | 82 (17.0 %) | 39 (10.0 %) |  |
| Philips | 273 (56.6 %) | 325 (83.3 %) |  |
| TOSHIBA (Canon Medical Systems) | 0 (0.00 %) | 1 (0.3 %) |  |

FLAIR=Fluid-attenuated inversion recovery; IQR=interquartile range. <sup>a</sup>Data were missing in a patient in the training dataset and in 5 patients in the external test dataset. <sup>b</sup>Data were missing in a patient in the external test dataset.

### Segmentation performance of deep learning algorithm

In the internal validation dataset (n=63), the model achieved a DSC of 0.531 (95% CI 0.497– 0.564) versus registered LA on CT scans (Table S2). Volumetric analysis showed that the predicted LA volume on CT correlated with registered LA volume on CT and WMH volume on FLAIR (ρ=0.898 and 0.813, respectively).

In the external test dataset, the DSC between predicted LA and registered LA on CT was 0.556 (0.545–0.566; Table 2). Representative cases with high DSC and low DSC between predicted LA and registered LA on CT in the external test dataset are shown in Figure 2. Volumetric analysis demonstrated excellent agreement between predicted LA volume and registered LA volume on CT (ρ=0.925; Figure 3A) and good agreement between predicted LA volume and WMH volume on FLAIR images (ρ=0.883; Figure 3A). With the increase of LA or WMH volumes, DSC increased in both internal validation and external test datasets (Figure S1). In 40 randomly selected cases with manual segmentation, the predicted LA volumes again demonstrated good agreement with manual segmentation (ρ=0.858; Figure S2). In addition, predicted LA volumes in the external testing dataset were strongly correlated with Fazekas grade (Pearson correlation coefficient=0.832; p<0.001; Figure 4A).

**Figure 2.**
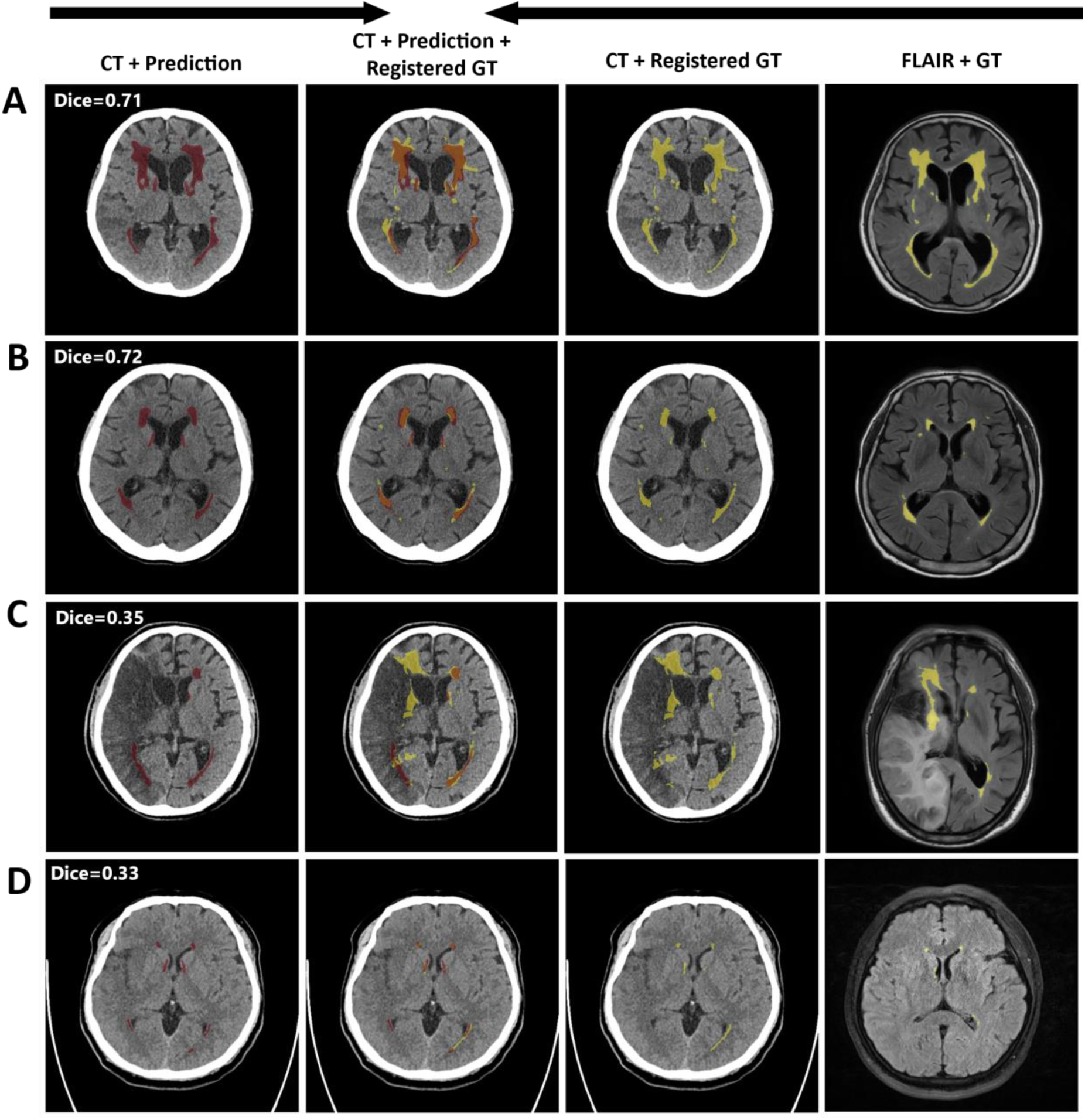
Representative cases demonstrating high and low Dice similarity coefficients (DSC) between predicted leukoaraiosis and registered leukoaraiosis from MR images in the external validation dataset. (A) High Dice, High WMH/Leukoaraiosis volume, (B) High Dice, Low WMH/Leukoaraiosis volume, (C) Low Dice, High WMH/Leukoaraiosis volume, (D) Low Dice, Low WMH/Leukoaraiosis volume.

**Figure 3.**
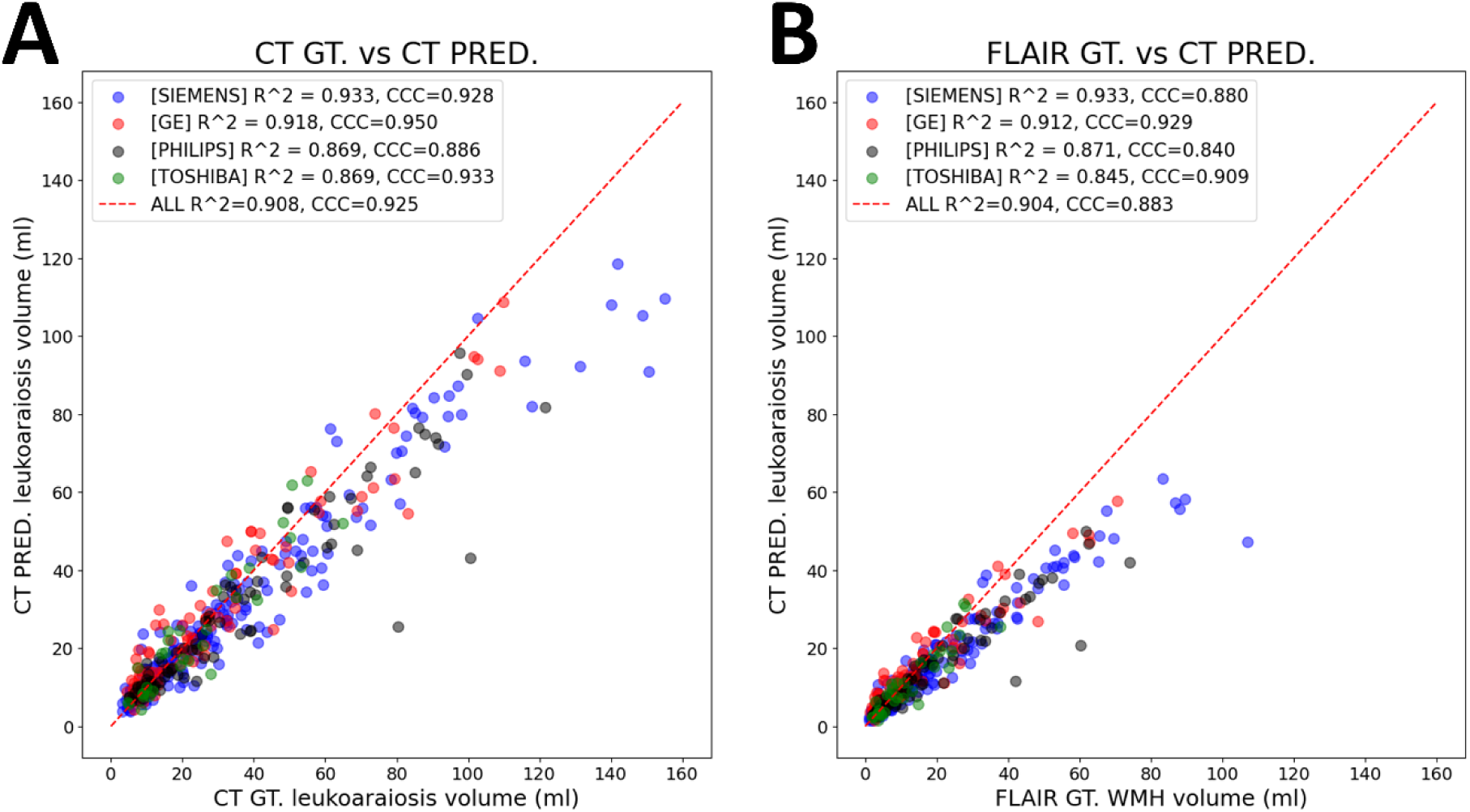
Volumetric correlation between automatically segmented leukoaraiosis volume on CT and ground truth on CT and MRI in the external test dataset. Dot plots showed a relationship between predicted versus registered leukoaraiosis volume (A) and between predicted leukoaraiosis volume on CT and predicted white matter hyperintensity volume on FLAIR (B). Each color represents a CT vendor. GT=ground truth; FLAIR=fluid-attenuated inversion recovery.

**Figure 4.**
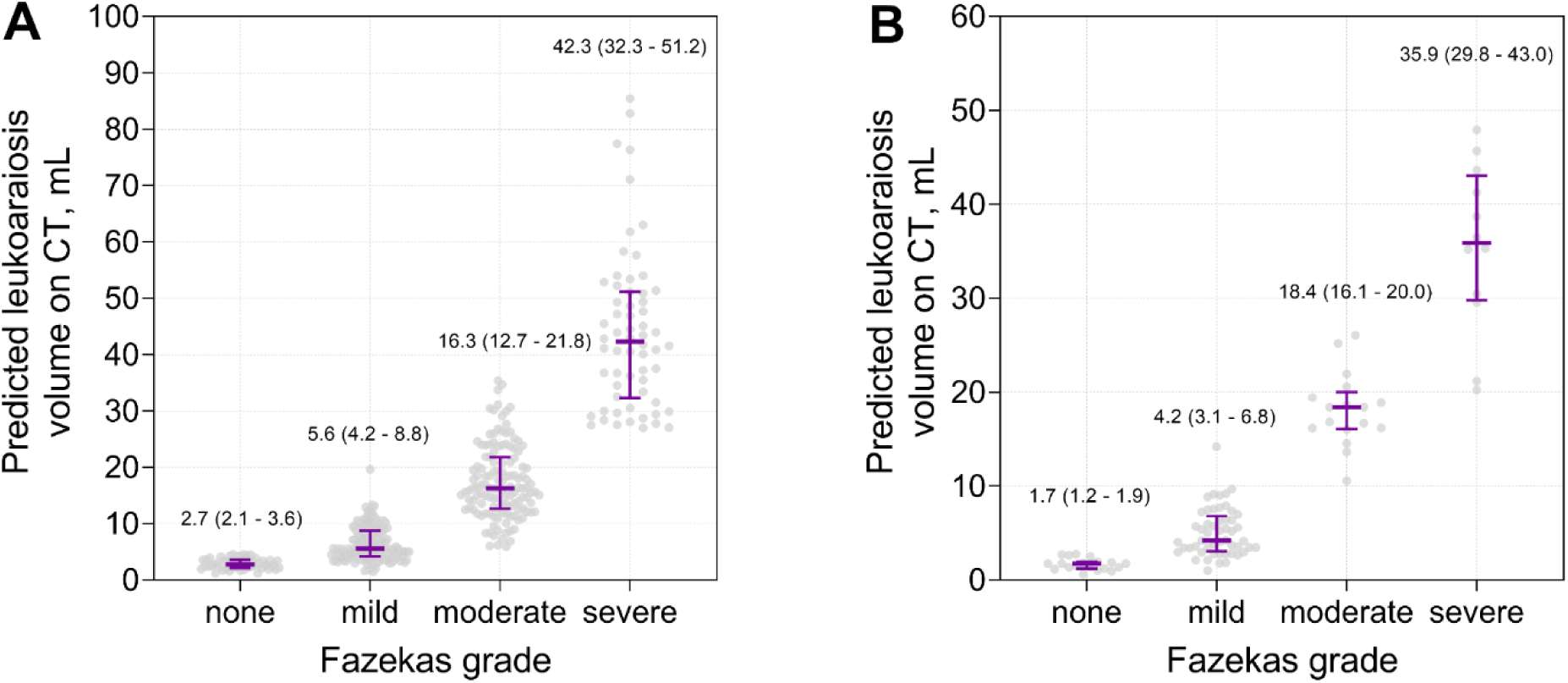
Volumetric correlation between the automatically segmented leukoaraiosis volume on CT and Fazekas grade in the external test dataset (A) and in the US population dataset (B). The numbers in the graph and purple lines (bars) indicate the median (interquartile range) of leukoaraiosis volumes for each Fazekas grade.

**Table 2.** Performance of deep learning algorithms segmenting leukoaraiosis on brain CT.

|  |  | All | SIEMENS | GE | PHILIPS | TOSHIBA |
| --- | --- | --- | --- | --- | --- | --- |
|  |  | 390 | 185<br>(47.4%) | 91<br>(22.1%) | 81<br>(20.8%) | 28<br>(7.17%) |
| CT<br>prediction<br>vs. CT GT | $r^2$ | 0.908<br>(0.890 - 0.926) | 0.933<br>(0.920 - 0.946) | 0.918<br>(0.886 - 0.949) | 0.869<br>(0.845 - 0.894) | 0.869<br>(0.845 - 0.894) |
|  | CCC | 0.925<br>(0.906 - 0.942) | 0.928<br>(0.914 - 0.941) | 0.950<br>(0.938 - 0.962) | 0.886<br>(0.819 - 0.952) | 0.933<br>(0.902 - 0.963) |
|  | DSC | 0.556<br>(0.545 - 0.566) | 0.564<br>(0.548 - 0.580) | 0.545<br>(0.524 - 0.566) | 0.558<br>(0.536 - 0.579) | 0.534<br>(0.493 - 0.574) |
| CT<br>prediction<br>vs. FLAIR<br>GT | $r^2$ | 0.904<br>(0.868 - 0.909) | 0.933<br>(0.920 - 0.946) | 0.912<br>(0.869 - 0.942) | 0.871<br>(0.846 - 0.895) | 0.845<br>(0.816 - 0.873) |
|  | CCC | 0.883<br>(0.856 - 0.908) | 0.880<br>(0.837 - 0.924) | 0.929<br>(0.907 - 0.951) | 0.840<br>(0.754 - 0.926) | 0.909<br>(0.830 - 0.988) |

Data were presented as estimate (95% confidence interval). GT=ground truth; FLAIR=fluid-attenuated inversion recovery; CCC=concordance correlation coefficient; DSC=Dice similarity coefficient.

In the US population (mean age 64.6±15.2 years [range: 24–90 years], 58.0% male), the predicted LA volumes showed a strong correlation with Fazekas grade (Pearson correlation coefficient=0.891; p<0.001; see Figure 4B).

### Subgroup analysis after stratification by CT vendors and infarct core volume on DWI

The model exhibited consistent segmentation performance independent of CT vendors (Table 2) with excellent agreement (CCC: 0.905–0.953). In comparison, the predicted LA volume and WMH on FLAIR showed moderate to substantial agreement (ρ ranging from 0.586 to 0.785).

In the external validation dataset, after stratification by infarct core volume (>10 mL [n=99] versus ≤10 mL [n=312]), the model showed excellent agreement with registered LA volume on CT in both groups (ρ=0.912 and ρ=0.922, respectively; Figure S3). In comparison to WMH volumes on FLAIR, the model exhibited good agreement (ρ=0.760 and ρ=0.786, respectively) in both groups.

### Clinical study using automatically measured LA volumes on CT

After exclusion, 867 consecutive patients with ischemic stroke were included in the clinical study. The mean age was 69.3 years (SD 13.0), and 39.2% were female (Table S3). The median predicted LA volume was 11.2 mL (IQR: 6.2–20.5 mL). Age was strongly associated with predicted LA volumes (coefficient 0.436, p<0.001; Figure S4). Multiple linear regression analysis showed that age, prior stroke, and atrial fibrillation were independently related to LA volumes (Table S4). Hypertension was independently associated with LA volumes in younger patients (<70 years) but not in elderly patients (≥70 years). In ordinal logistic regression analysis, the third, fourth, and fifth quintiles of LA volumes were incrementally associated with higher 3-month mRS scores, respectively (Table 3). After adjusting for covariates, the association between LA quintiles and mRS scores was slightly attenuated but remained significant, with adjusted odds ratios of 1.59 (95% CI: 1.08–2.36) and 1.65 (95% CI: 1.10–2.46) for the fourth and fifth quintiles, respectively.

**Table 3.** Univariate and multivariable ordinal logistic regression analysis between quintiles of leukoaraiosis volumes and modified Rankin Scale score at 3-months.

|  | Crude odds ratio (95% CI) | P value | Adjusted<sup>a</sup> odds ratio (95% CI) | P value |
| --- | --- | --- | --- | --- |
| 1st quintile | Reference |  | Reference |  |
| 2nd quintile | 1.35 (0.92 – 1.96) | 0.12 | 1.16 (0.78 – 1.71) | 0.46 |
| 3rd quintile | 1.59 (1.09 – 2.31) | 0.015 | 1.49 (1.01 – 2.19) | 0.045 |
| 4th quintile | 1.77 (1.21 – 2.58) | 0.003 | 1.59 (1.08 – 2.36) | 0.02 |
| 5th quintile | 2.14 (1.46 – 3.12) | < 0.001 | 1.64 (1.10 – 2.46) | 0.016 |

<sup>a</sup>Adjusted for age, admission National Institutes of Health Stroke Scale score, sex, body mass index, hypertension, diabetes, hyperlipidemia, smoking, atrial fibrillation, coronary artery disease, and revascularization therapy.

## Discussion

In the present study, a deep learning algorithm that automatically segments LA on head CT exams was developed using a CT-MRI_FLAIR_ paired dataset without human annotation and externally validated in an independent international (Korea and US), multicenter, multi-vendor dataset. The predicted LA volumes on CT exhibited excellent agreement with WMH volumes on MRI across multiple CT vendors showing generalizability. The predicted LA segmentations correlated well with manual segmentations outlined by an expert and a visual rating scale in both external testing and US datasets. Using a third clinical dataset, we show the predicted LA volumes are indeed associated with vascular risk factors and stroke outcome. Several studies have reported on deep learning algorithms for segmenting LA on CT scans.^9,25,26^ Chen et al.^9^ demonstrated that the automated LA volume correlation at MRI was 0.85 and at CT imaging was 0.71 when compared with LA volumes segmented by experts, which is lower compared to our results. Pitkanen et al.^26^ developed a convolutional neural network algorithm using 147 paired CT-MRI_FLAIR_ images and reported a volumetric correlation of 0.94. However, they validated the algorithm using the same training data. Voorst, et al.^25^ developed an algorithm using 245 CT exams with expert annotations and reported a DSC of 0.68. However, the performance of the algorithm performed poorly in external validation testing, with a DSC of 0.23. Our algorithm, trained on a large CT-MRI_FLAIR_ paired dataset, exhibited robust performance on an external dataset with DSC ranging from 0.54 to 0.60, and represents high performance for an externally verified LA segmentation algorithm.

Visual scoring systems for LA on CT, despite being widely used, are limited by their reliance on subjective visual criteria and the resultant variability in interrater reliability.^8,27^ This variability can hinder accurate diagnosis and monitoring of disease progression. In contrast, the deep learning algorithm developed in this study offers objectivity and reproducibility. By eliminating human subjectivity, the algorithm enhances diagnostic accuracy and provides a reliable tool for assessing LA. This advancement is particularly important for large-scale studies and clinical trials where reproducibility in LA measurement is crucial.

A critical aspect of the deep learning algorithm’s validation involved testing its performance across multiple CT vendors. The algorithm demonstrated high CCC values, ranging from 0.905 to 0.953, indicating excellent cross-vendor agreement. Consistent performance across different imaging vendors ensures that the algorithm can be widely adopted and provide reliable LA segmentations. This eliminates the need for image harmonization within and across institutions. This universality is a significant step towards standardizing LA assessment in clinical practice.

In the external testing dataset, the predicted LA volumes on CT significantly correlated with WMH volumes on FLAIR MRI. This correlation is crucial as it validates the algorithm’s effectiveness in translating the more precise measurements typically obtained from MRI into the more commonly available CT scans.^28^ In addition, CT scans remain the primary modality for patients presenting with neurological symptoms although MRI is superior to CT in the diagnosis of stroke.^29^ The ability to accurately assess LA on CT, using an algorithm that correlates well with MRI-derived volumes, bridges the gap between the two imaging modalities. A strong correlation between predicted LA volumes and Fazekas grade in both Korean and US populations further supported the generalizability of our algorithm. Furthermore, using an independent clinical dataset, we demonstrated associations between automatically measured LA volume on CT and both risk factors and clinical outcomes after ischemic stroke, consistent with the known literature in studies using FLAIR MRI.^2,3^ These results bolster the reliability and reproducibility of our algorithm, enhancing patient management where MRI is not readily available.^30^

In the present study, we developed an algorithm for segmenting LA on CT without human annotation. By utilizing CT-MRI_FLAIR_ paired data, the algorithm eliminates the need for labor-intensive manual annotations, thereby streamlining the segmentation process. This approach ensures a consistent and objective analysis, free from the variability and potential biases inherent in human annotations in outlining obscure LA on CT.^31–33^ The ability to accurately segment LA on CT without human intervention enhances the algorithm’s efficiency and reliability, making it a valuable tool for clinical practice and large-scale studies.

### Limitations

Although the algorithm was validated using multicenter, multi-vendor data, the training data was limited to Asian patients with ischemic stroke. However, in the US population, we observed a strong correlation between predicted LA volumes and Fazekas grade, indicating that our algorithm may be effective across different racial groups. Additionally, the exclusion criteria applied to the initial patient cohort, particularly the exclusion of patients with more than 5 mL of ischemic stroke on DWI, may have introduced bias into the training dataset. Nonetheless, subgroup analysis showed that the algorithm maintained its performance in patients with large infarcts on DWI, albeit with slightly lower accuracy compared to those with smaller infarcts. Even though our algorithm demonstrated a strong volume correlation between predicted LA volumes on CT and WMH volumes on FLAIR, regional similarity, as assessed by DSC, was relatively low. Weak DSC can be explained by the limitations of co-registering different imaging modalities,^34^ which limits the usability of our algorithm in studies where spatial correlation is crucial.

## Conclusion

By providing a more accurate, reproducible, and accessible method for assessing LA, the proposed algorithm has the potential to improve patient care and outcomes. Its robustness across different imaging systems and validated correlation with MRI-derived volumes enhances its clinical utility, making it a valuable tool for both routine clinical practice and research.

## Data Availability

Data will be made available upon request to the corresponding author.

## Acknowledgment

The authors appreciate the contributions of all members of the Clinical Research Collaboration for Stroke in Korea to this study.

## Sources of Funding

This research was supported by the Multiministry Grant for Medical Device Development (KMDF_PR_20200901_0098), funded by the Korean government and a grant of the Korea Health Technology R&D Project through the Korea Health Industry Development Institute, funded by the Ministry of Health & Welfare, Republic of Korea (grant number: HI22C0454).

## Disclosure

Wi-Sun Ryu, Ju Hyung Lee, Dongmin Kim, and Myungjae Lee are employees of JLK Inc., Republic of Korea.

## Non-standard Abbreviations and Acronyms

WMH: white matter hyperintensity

LA: leukoaraiosis

DSC: Dice similarity coefficient

FLAIR: Fluid-attenuated inversion recovery

mRS: modified Rankin Scale

NIHSS: National Institute of Health Stroke Scale

HU: Hounsfield Unit

ReLU: rectified linear unit

CCC: Concordance Correlation Coefficient

## Notes

### Author Declarations

The study protocol was approved by the institutional review board of Seoul National University Bundang Hospital (B-2307-841-303) and all subjects or their legal proxies provided a written informed consent.

